# Is the psychological well-being of a population associated with COVID-19 related survival?

**DOI:** 10.1101/2020.06.05.20123018

**Authors:** Frederik Feys

**Affiliations:** PhD Medical Sciences, Independent researcher, Antwerp, Belgium

**Author notes:** Funding: None.

**Keywords:** public mental health, COVID-19, well-being, mortality, risk factor

## Abstract

**OBJECTIVE:** Immunological mind-body research suggests mental health may also be important in the COVID-19 pandemic. This study aimed to investigate the potential influence of mental health as a protective factor for COVID-19 related mortality in the general population. The second goal was to examine this among populations of countries most affected by COVID-19 related mortality.

**METHODS:** Data sources were the Global Burden of Disease report 2017 and publicly reported situational reports of COVID-19. We described variables; calculated the spearman’s correlation coefficient, calculated the percentage of the variability of the data that is explained by the association. We explored inter-relationships among other variables: aged 70 or older, cardiovascular disease, obesity and diabetes. A correlation matrix with plotted scatter matrix diagrams was produced.

**RESULTS:** Across 181 countries, the mean total COVID-19 related survivors per million was 999,949 (sd = 125), median = 999,993. The variable had a lognormal distribution; the mean mentally healthy per 100,000 was 85,411 (sd = 1,871), median = 85,634. The test of normality resulted in p-value < 0.001. Correlation of mentally healthy per 100,0000 and totals of COVID-19 related survivors was *ϱ*_s_ = 0.29 (n = 181, 95% CI 0.16–0.43). The variance explained by the relation between mental healthy and totals of COVID-19 related survivors was 8.4% (2.6–18.5%). Across countries most affected by COVID-19 related mortality *ϱ*_s_ = 0.49 (n = 45, 0.28–0.70), explaining 24.2% (7.7–49.3%).

**CONCLUSION:** A weak association was found between the psychological well-being of a population and COVID-19 related survival. This relationship explained between 2.6 and 18.5% of COVID-19 related survival. For countries most affected by COVID-19 related death, this association was moderate and explained between 7.7 and 49.3%. Confirmation of these important observational findings is needed with future individual patient data research.

## Introduction

The connection between body and mind shows well in the placebo effect (1). In response to a placebo, positive expectations can lead to better health outcomes. Conversely, there will be a nocebo effect if negative expectations lead to harms (2). For example, fake surgery in orthopedics can relieve the pain just as much as its “real” variant (3). It is important that those studies are conducted double-blind: neither the researchers nor the participants know who gets which treatment. Clinical studies with an informed consent that contains too much information about possible disadvantages can undermine the double-blind nature of the study. Participants who discover that they are receiving placebo have lower expectations for improvement (4).

In infectious diseases, such as after infection with the new SARS-CoV-2 virus, the body reacts with an immune response. The extent to which this happens successfully, determines its cure. The wisdom *Mens sana in corpore sano* implies that a healthy mind also drives a better immune response. There is a lot of evidence for this immunological mind-body connection; psychological stress was associated in a dose-response manner with an increased risk of acute infectious respiratory illness (5). Psychological well-being led to better survival in patients with cardiovascular disease or after infection by the human immunodeficiency virus (6). More than 30 years of research on psychoimmunology shows that the mind influences all important immunological parameters (7). It is therefore no surprise that vaccines work more effectively in psychologically healthy people (8).

A very large sampled UK study showed that COVID-19 related deaths are associated with various risk factors: aged 70 or older, poverty, immunosuppressive conditions, being male, diabetes, obesity, and cardiovascular disease (9). Unfortunately, mental health factors such as anxiety, depression or substance use were not investigated. This leaves a gap in our knowledge.

Worldwide it appears that some countries are more affected by COVID-19 related mortality. Epidemiological research refers to all kinds of factors, such as differences in the age structure of the population (10), the health care capacity(11) and cultural differences. The possible importance of mental health in more affected countries remains unclear.

This research aims to investigate the potential influence of mental health as a protective factor for COVID-19 related mortality in the general population. A second aim is to verify this in the countries most affected by COVID-19 related mortality. For the completeness and estimation of the relative importance, we incorporated some important risk factors associated with COVID-19 related survival such as older age, diabetes, cardiovascular disease and obesity.

## Methods

A mentally healthy (MH) population is defined as the proportion of a countries population free from mental or substance use disorders. Along with other risk 5 factors (cardiovascular disease, obesity, diabetes), the Global Burden of Disease report 2017 provided the data (12). Because age stratification differs across countries, age-standardized estimates were taken. Totals of COVID-19 related survival (TCS) was estimated from publicly reported situational reports of countries as of June, 9th 2020 (13). The final dataset included any country with data on psychological well being.

### Statistical analysis

We calculated means and standard deviations for every variable, performed Shapiro and Wilk’s W test to assess normal distributions, plotted histograms and normal plots (see plots in Appendix 1). We calculated the correlation coefficient *ϱ* to measure the degree of association, along with its p-value. A 95% CI was calculated for the correlation of the main variables. Variables were not normally distributed, so we used Spearman’s rank correlation. It also allowed us to assess not linear association but a more general association. We calculated 100*ϱ*^2^, the percentage of the variability of the data that is explained by the association of the two main variables.

We explored inter-relationships among other variables too: aged 70 or older, cardiovascular disease, obesity and diabetes. A correlation matrix stated the *ϱ* value along with the number of observations. We plotted scatter matrix diagrams to visually inform an underlying trend.

#### Effect modification

The group of countries most affected consisted of 25% (quartile) of all countries with the lowest COVID-19 related survival rates. Correlations were calculated with additional risk factors. Depending on the number of cases, additional risk factors could be included in the analysis. The criterion was at least ten observations (i.e. ten countries in a subgroup) should be available for each characteristic. Additional considered characteristics in order of importance (in line with UK OpenSafely study): aged 70 or older, diabetes, obesity, and cardiovascular disease.

## Results

### Descriptives

Across 181 countries, the mean total COVID-19 related survivors per million was 999,949 (sd = 125), median = 999,993. The test of normality resulted in p-value < 0.001; the mean mentally healthy per 100,000 was 85,411 (sd = 1,871), median = 85,634. The test of normality resulted in p-value < 0.001 (Table 1).

**Table 1.** Descriptives for total COVID-19 related survivors; mentally healthy rates and 4 explanatory variables

|  |  |  |  |  |  |  |
| --- | --- | --- | --- | --- | --- | --- |
| variable |  |  |  |  | Shapiro and Wilk's W test |  |
|  | n | mean | sd | median | W | p-value |
| total COVID-19 related survivors per million | 181 | 999,949 | 125 | 999,993 | 0.444 | <0.001 |
| mentally healthy per 100,000 | 181 | 85,411 | 1,871 | 85,634 | 0.945 | <0.001 |
| aged 70 or older percent | 175 | 5.50 | 4.25 | 3.83 | 0.858 | <0.001 |
| diabetes per 100,000 | 137 | 7,372 | 2,438 | 6,738 | 0.883 | <0.001 |
| obesity percent | 176 | 18.15 | 8.90 | 20.20 | 0.946 | <0.001 |
| CVD per 100,000 | 181 | 6,312 | 766 | 6,200 | 0.969 | <0.001 |

### Correlation

Correlation of mentally healthy per 100,0000 and totals of COVID-19 related survivors was *ϱ*_s_ = 0.29 (n = 181, 95% CI 0.15–0.43) (Table 2). The variance explained by the relation between mental healthy and totals of COVID-19 related survivors was 8.4% (95% CI 2.6 –18.5).

**Table 2.** Correlation matrix with *ϱ*_s_ for total COVID-19 related survivors; mentally healthy rates and 4 explanatory variables

| variable | TCS | MH | Age≥70 | Diabetes | Obesity |
| --- | --- | --- | --- | --- | --- |
| MH | 0.29***<br>(0.15-0.43)¥ |  |  |  |  |
| Age≥70 | -0.55*** | -0.17* | | | |
| Diabetes | 0.02 | 0.01 | -0.10 |  |  |
| Obesity | -0.46*** | -0.42*** | 0.42*** | 0.20** |  |
| CVD | -0.22*** | -0.18* | 0.16* | 0.31*** | 0.38*** |
| abbreviations: TCS: total COVID-19 related survivors, CVD: cardiovascular disease, MH: mentally healthy |  |  |  |  |  |
| *** p-value < 0.001, ** p-value < 0.01, * p-value < 0.05 |  |  |  |  |  |
| ¥ 95% confidence interval |  |  |  |  |  |

**Figure 1.**
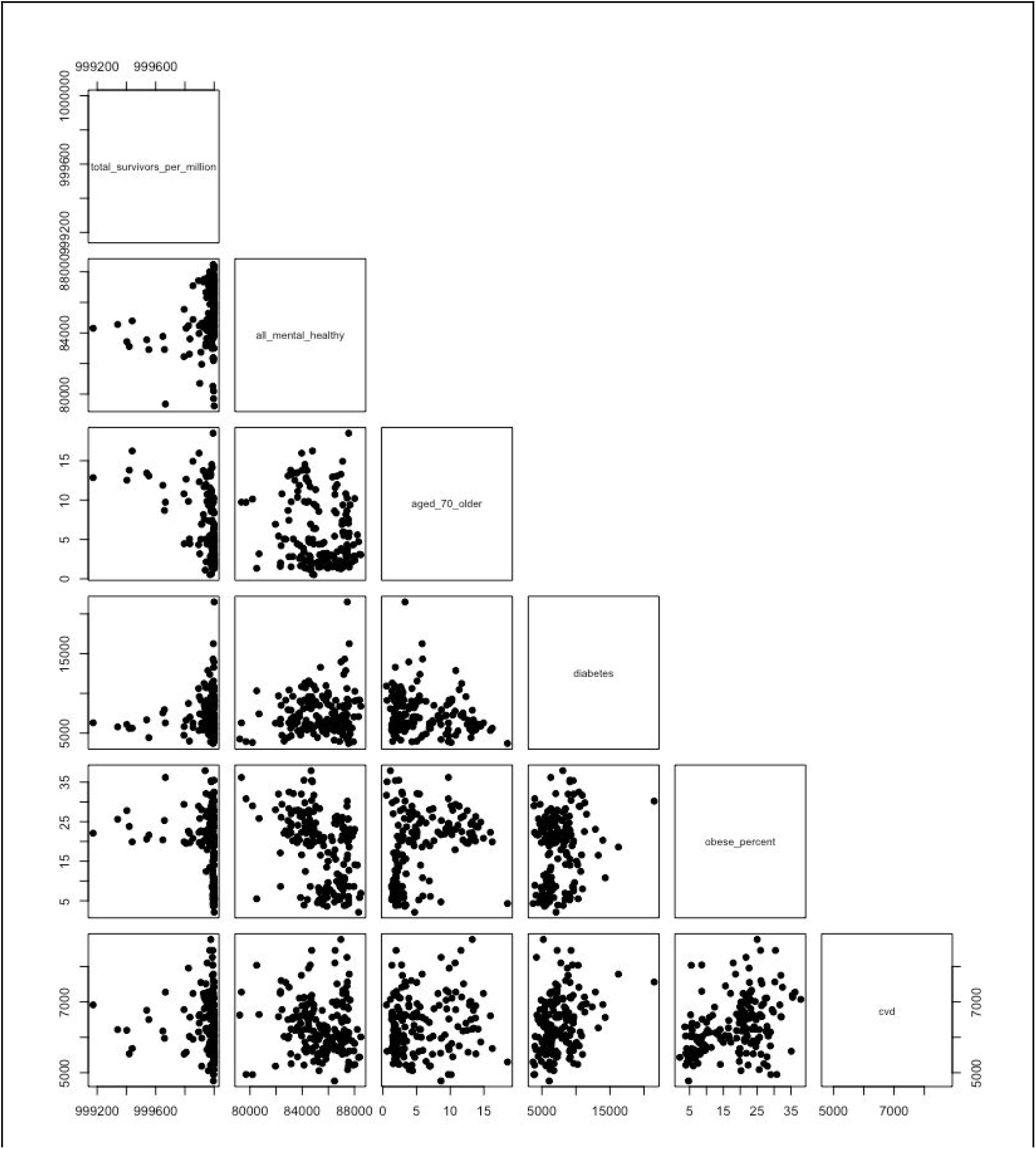
Scatter diagrams corresponding to Table 2

### Effect modification

Descriptives and scatter diagrams for the group of 45 countries most affected by COVID-19 related mortality can be found in Appendix 2.

**Table 3.** Correlation matrix with *ϱ*_s_ for total COVID-19 related survivors; mentally healthy rates and 3 explanatory variables

| variable | TCS | MH | Age≥70 | Diabetes |
| --- | --- | --- | --- | --- |
| MH | 0.49***<br>(0.28-0.70)¥ |  |  |  |
| Age≥70 | -0.34* | -0.04 |  |  |
| Diabetes | 0.24 | 0.13 | -0.33* |  |
| Obesity | -0.04 | -0.21 | -0.22 | -0.03 |
| abbreviations: TCS: total COVID-19 related survivors, MH: mentally healthy<br>*** p-value < 0.001, ** p-value < 0.01, * p-value < 0.05<br>¥ 95% confidence interval |  |  |  |  |

## Discussion

### Summary

Across 181 countries, mentally healthy populations showed a weak association with COVID-19 related survival. Mentally healthy populations explained 8.4% (95% CI 2.6 –18.5) of COVID-19 related survival.

In line with other research (in decreasing order); populations aged 70 or older, obesity and cardiovascular disease were the drivers worsening COVID-19 related survival. Our results did not confirm that diabetes is a risk factor (9,15,16).

We found a weak to moderate association between mentally healthy populations and lower rates of obesity. This finding confirms results from a 13 countries survey, that found statistically significant obesity–mental disorder associations (17).

The secondary analysis with the 45 countries most affected with COVID-19 related death shows a moderate relationship *ϱ*_s_ = 0.49 (0.28–0.70) between mental well-being and COVID-19 related survival.

### Strengths and weaknesses

This is the first study to assess the potential association between mental healthy populations and their chance of COVID-19 related survival. Thanks to the Global Burden of Disease study, we could cover mental health of 181 (93%) countries worldwide. All analyses were underpinned by scientific rationale and were performed using appropriate statistical methods.

The study’s internal validity is limited. The countries in the dataset do not come from a random sample. The reported p-values and confidence intervals are therefore not strictly valid, but give an idea of the uncertainty surrounding the values found. Mental health data stemmed from 2017, so they do not describe prevalences today. However, we assumed that prevalences for mental disorders and substance use have remained fairly constant up to this day in most countries and therefore remain a valuable proxy for the current mental health rates. Still, the COVID-19 pandemic has lowered mentally healthy populations in some countries (18–20); but exactly how more data from other countries would alter our findings remains to discover. Attribution of death to a specific cause is often challenging and definitions of “COVID-19 death” vary across countries and sometimes even change within countries over time. Overall, some COVID-19 deaths may be missed, and others may be overcounted.

Clearly, the external validity of findings to individual countries is limited. Findings are observational and should be interpreted cautiously because of the risk of confounding.

### Policy Implications and Interpretation

Many countries have policies that focus solely on the viral components of the COVID-19 pandemic. The emphasis lies on strictly physical factors in the prevention and approach. However, we were able to demonstrate that there is a weak link between psychologically healthy populations and a protection against COVID-19 related death. For countries most affected by COVID-19 related death, we found a moderate link between psychological well-being and COVID-19 related survival.

It is too early to make interventions based on this study only. However, the prudence principle applies: it is important to give preventive attention to a mentally healthy population in order to strengthen the chances of survival after viral contamination. Particular attention also seems to be needed for the overweight population. We found that they experience reduced mental health. A synergistic effect could occur so that their chances of COVID-19 related survival further decrease. 1 in 7 people on this planet suffer from poor mental health or substance use and its association with covid-19 related survival makes improved mental health strategies urgently needed.

The unexpected finding that the psychological well-being in countries most affected by COVID-19 related mortality appears to have an impact on the survival after SARS-CoV-2 virus infection makes populations suffering from mental illness and substance use at risk for COVID-19 related mortality.

### Future research

Government agencies have the responsibility to make the large amounts of data, especially through primary care, available for research communities to better tackle the COVID-19 crisis. Doing so, individual patient data study would allow more valid and –if done on a large scale- also more precisely estimate the impact of mental health on COVID-19 related survival.

### Conclusion

Across 181 countries, a weak association was found between mentally healthy populations and COVID-19 related survival. This relationship explained between 2.6 and 18.5% of COVID-19 related survival. For countries most affected by COVID-19 related death, this association was moderate and explained between 7.7 and 49.3% of COVID-19 related survival. Sharing of patient mental health data is urgently needed and would allow better understanding of the psychoimmunology of infectious diseases.

## Data Availability

data available on ResearchGate:
10.13140/RG.2.2.27492.40323

## History

| Date | Event | Comment |
| --- | --- | --- |
| 9/6/2020 | updated with recent data (June 8th, 2020).<br>Reported a secondary analysis of countries most affected by covid-19 mortality. |  |
| 5/6/2020 | preprint manuscript version 1 submitted to medrxiv | accepted June 7th, 2020 |

## APPENDIX 1 Main analysis: all countries

### Histograms and normality plots

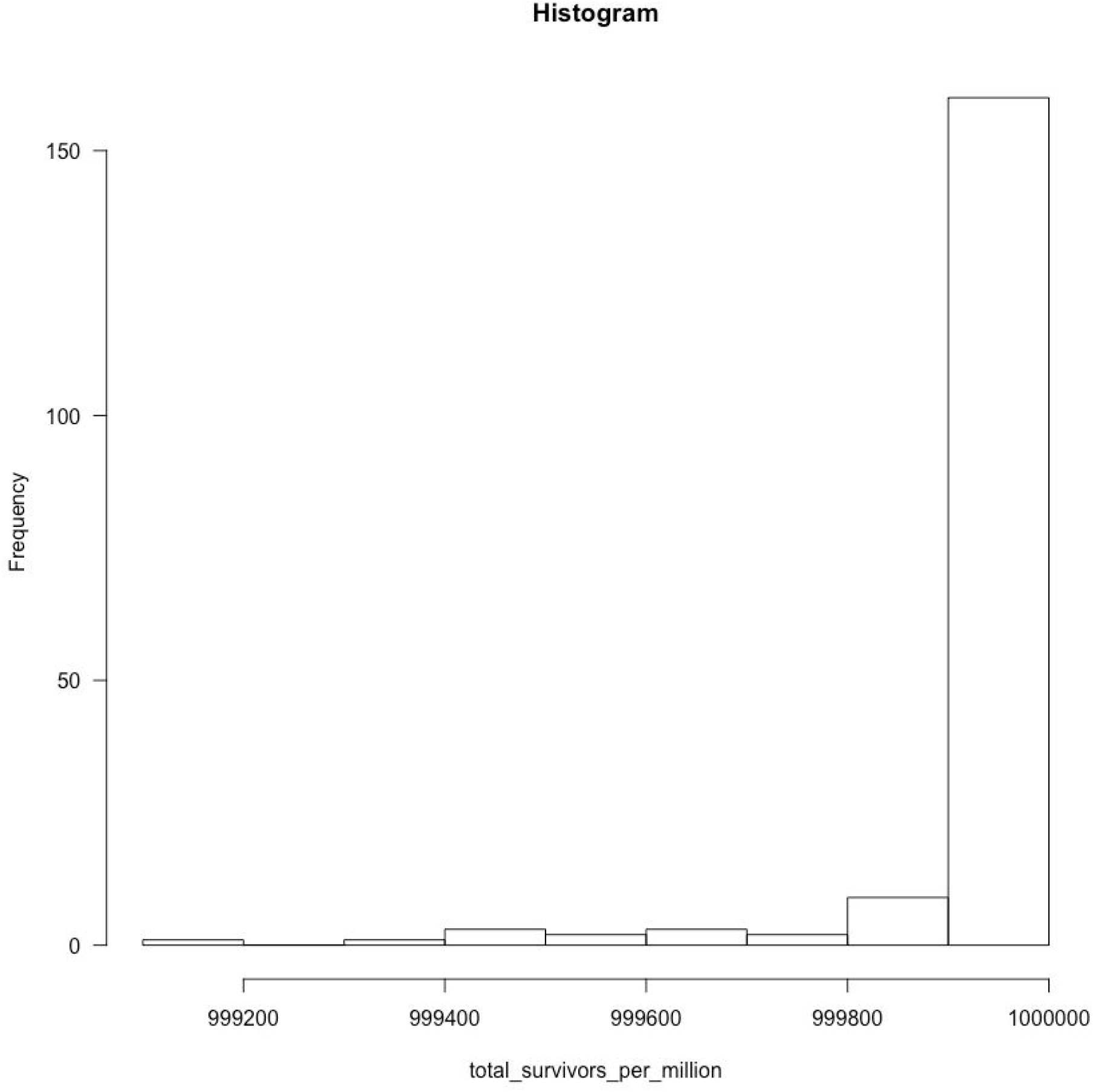

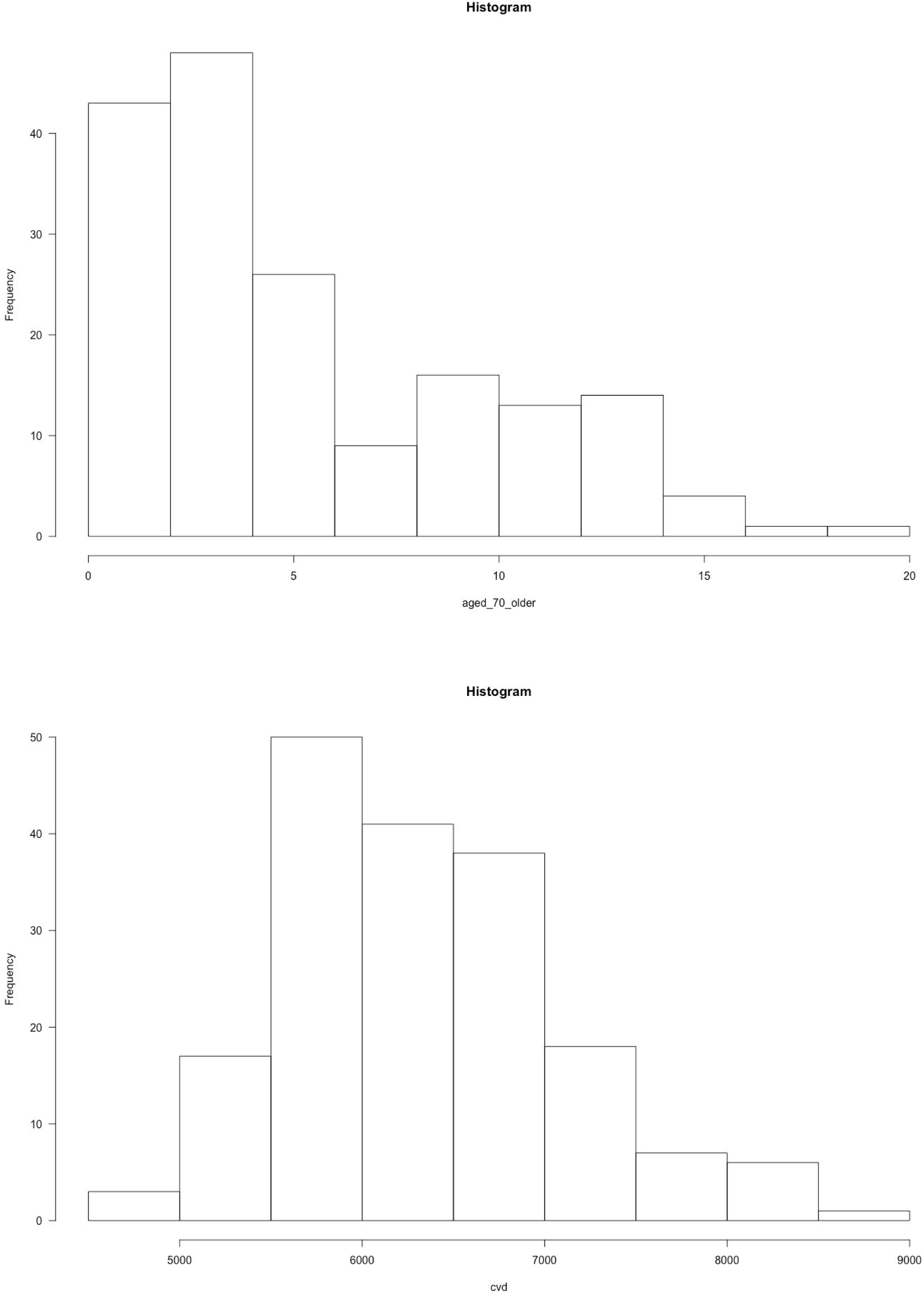

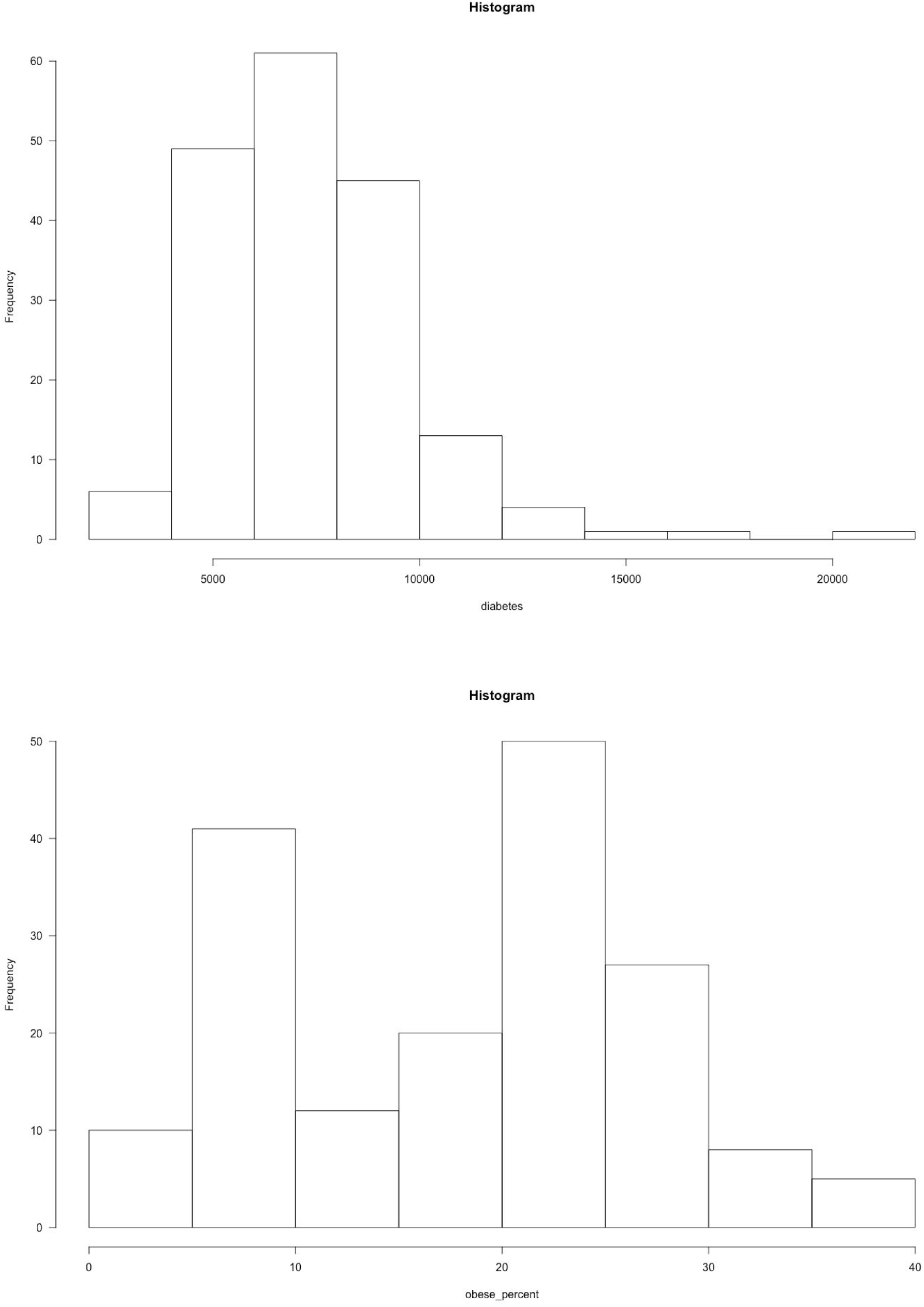

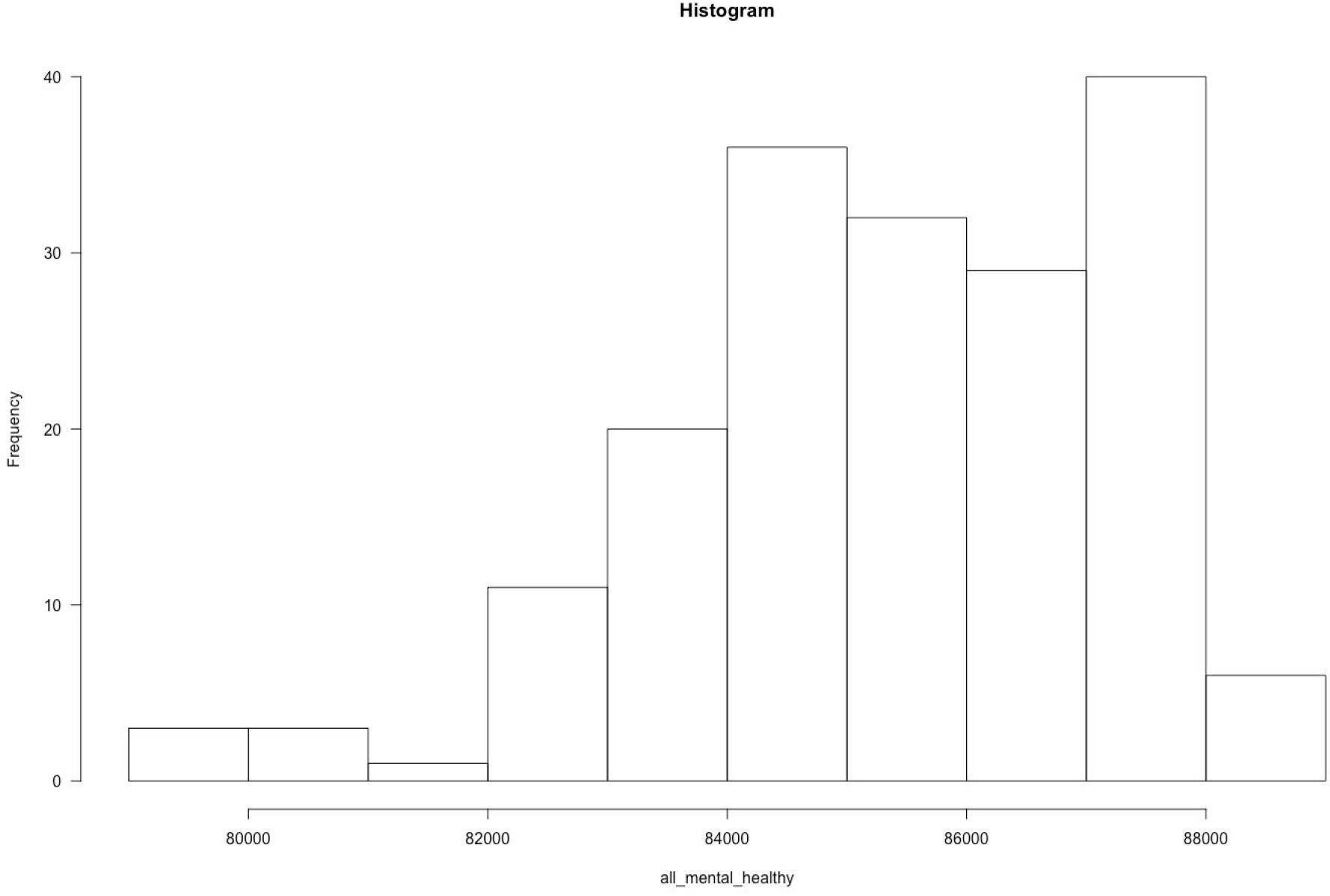

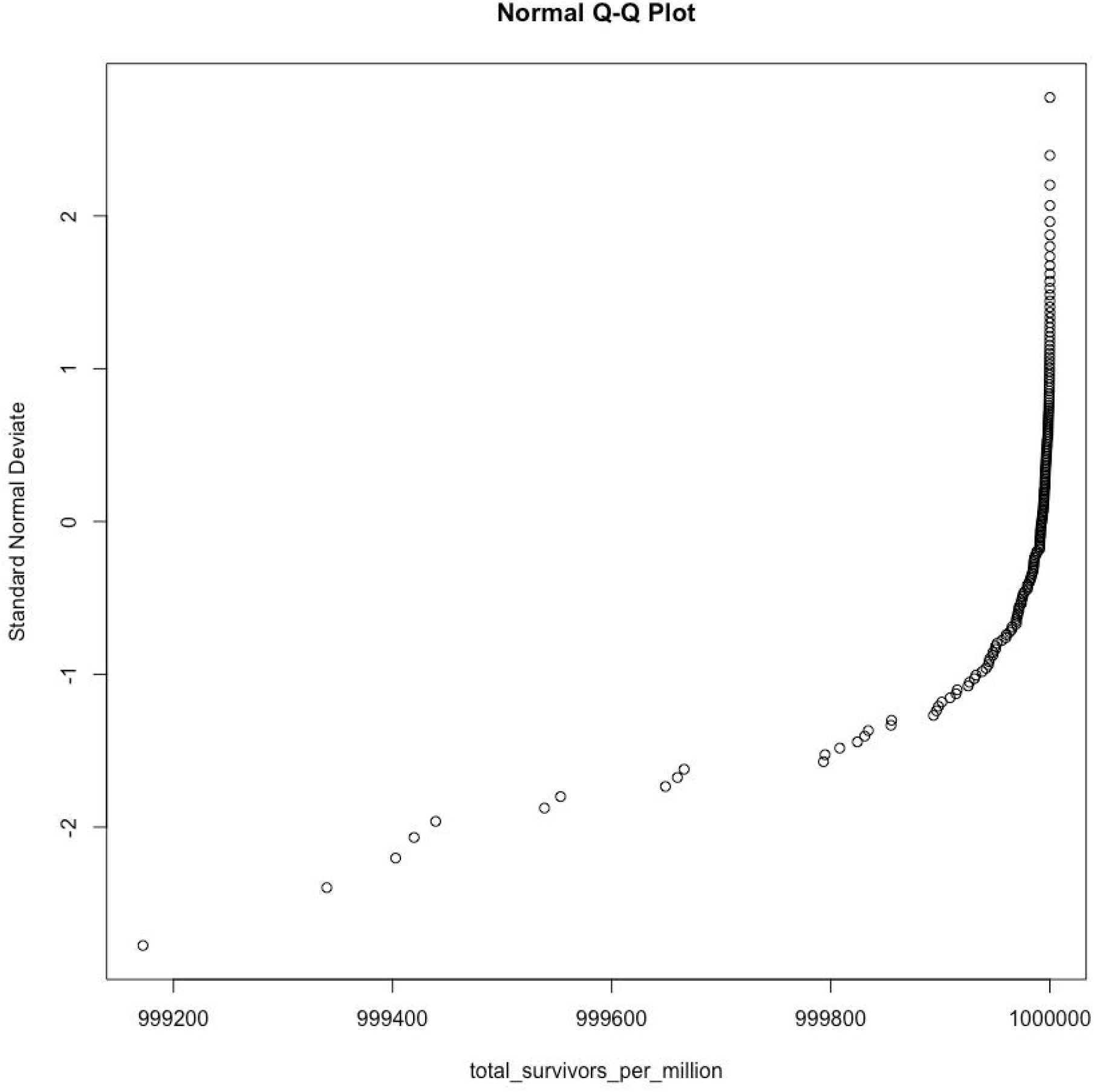

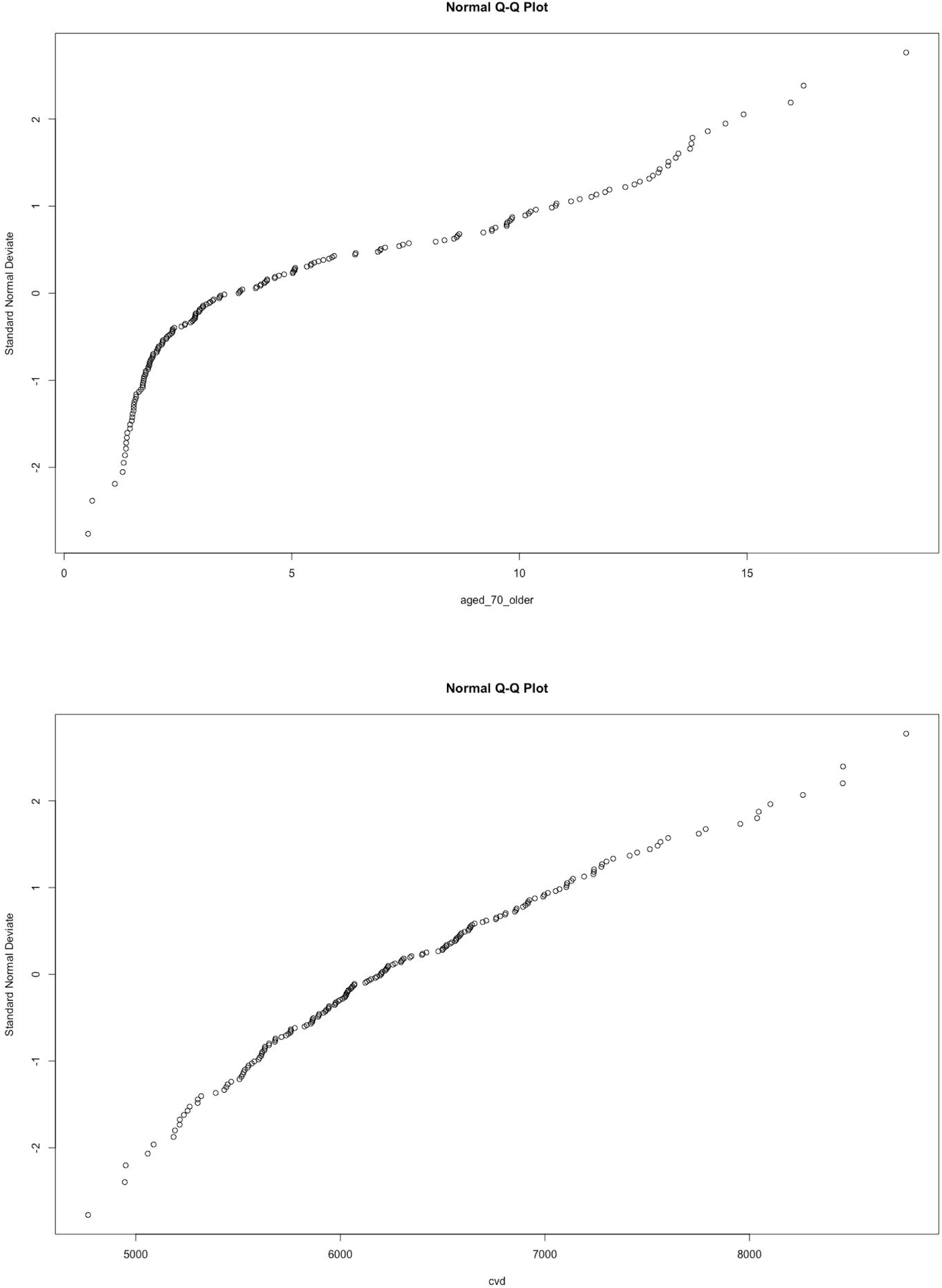

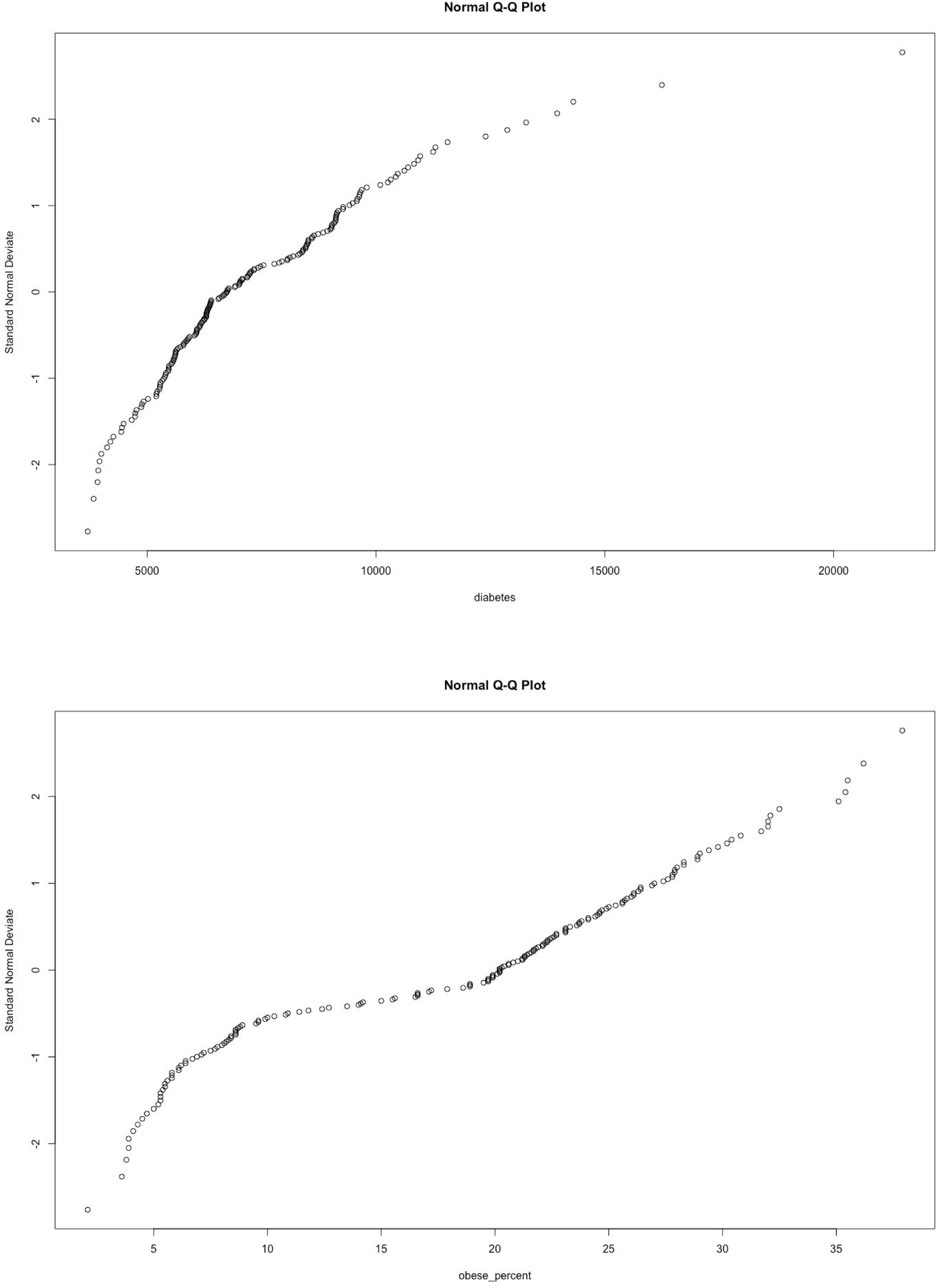

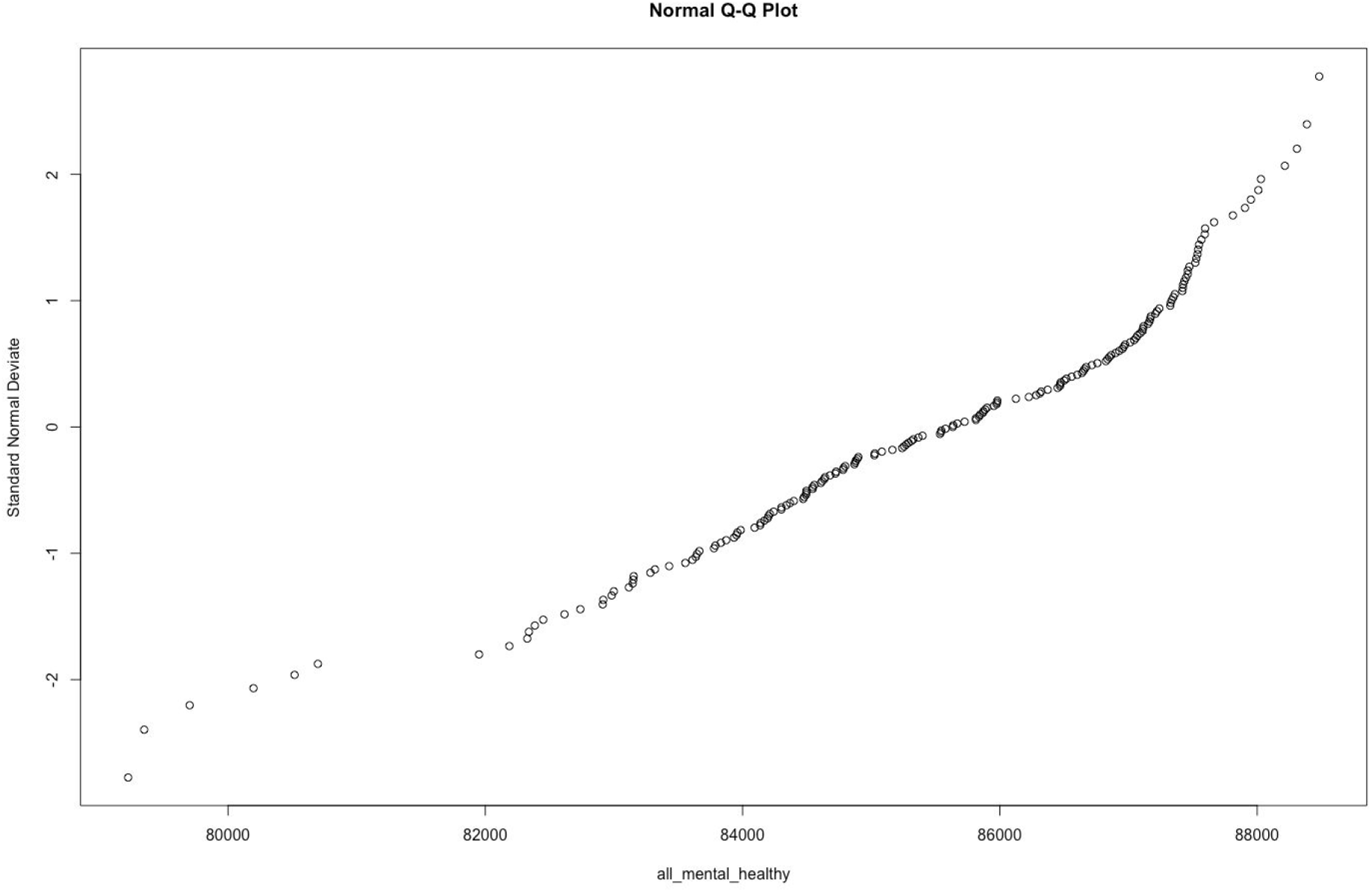

## APPENDIX 2 Secondary analysis: countries most affected by COVID-19 related mortality

### Descriptives

**Table 3.**
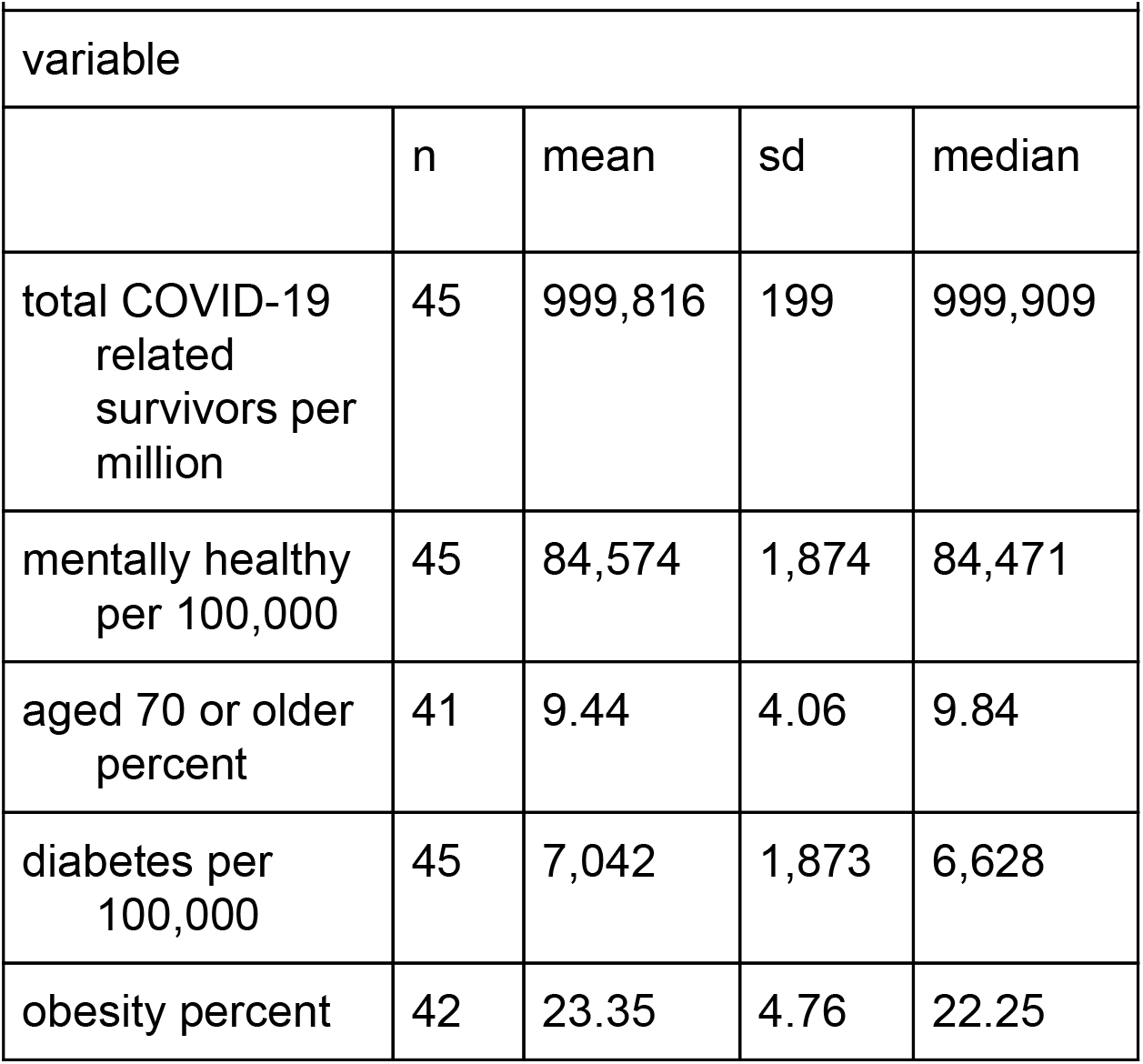
Descriptives for total COVID-19 related survivors; mentally healthy rates and 4 explanatory variables

|  |  |  |  |  |
| --- | --- | --- | --- | --- |
| variable |  |  |  |  |
|  | n | mean | sd | median |
| total COVID-19 related survivors per million | 45 | 999,816 | 199 | 999,909 |
| mentally healthy per 100,000 | 45 | 84,574 | 1,874 | 84,471 |
| aged 70 or older percent | 41 | 9.44 | 4.06 | 9.84 |
| diabetes per 100,000 | 45 | 7,042 | 1,873 | 6,628 |
| obesity percent | 42 | 23.35 | 4.76 | 22.25 |

### Scatter diagrams

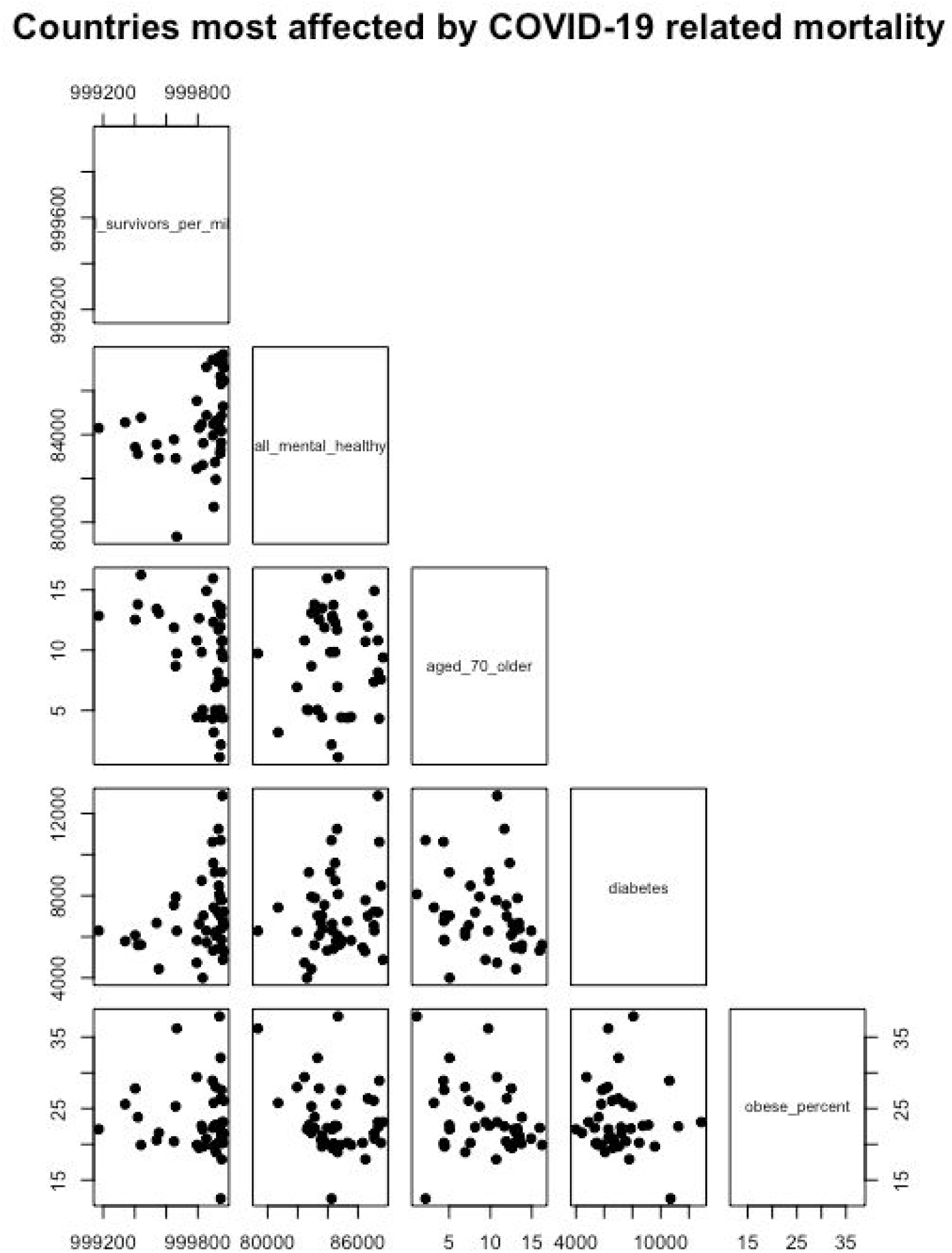

